# Structural and Functional Connectomic Signatures of Durable Tremor Control After MRgFUS Thalamotomy in Parkinson’s Disease

**DOI:** 10.64898/2026.03.31.26349811

**Authors:** Jonas Krauss, Neeraj Upadhyay, Marcel Daamen, Veronika Purrer, Valeri Borger, Hannah Weiland, Lisa Steffens, Alexander Radbruch, Markus Essler, Julian Luetkens, Ullrich Wüllner, Henning Boecker

## Abstract

Magnetic resonance-guided focused ultrasound (MRgFUS) thalamotomy is an established thermoablative treatment for tremor. Although outcomes in Essential Tremor approach those of deep brain stimulation, efficacy in tremor-dominant Parkinson’s disease (TDPD) is often less durable, with tremor relapse reported in 30-50% of cases. Previous associations with lesion size or age remain descriptive and do not explain why anatomically similar lesions yield divergent long-term outcomes.

We retrospectively analyzed 20 patients with TDPD who underwent unilateral MRgFUS. Lesions were used as seeds for normative structural and functional connectivity analyses.

Durable tremor control was associated with lesion showing stronger functional connectivity to primary motor (M1), primary somatosensory (S1), and supplementary motor areas, as well as inferior frontal and occipital cortices. In contrast, relapse was linked to greater connectivity with cerebellar motor and associative regions. Structurally, optimal lesions converged at the triangular interface of the nuclei ventralis intermedius, ventralis oralis, and ventro caudalis. Streamlines associated with better outcomes projected posteriorly towards S1, with M1 delineating an anterior functional boundary beyond which outcomes declined. Structural fingerprints emphasized posterior sensorimotor areas as critical therapeutic outputs.

Findings define a connectivity-based substrate of durable tremor suppression and support the development of individualized, network-guided targeting strategies for MRgFUS in TDPD.

## Introduction

Magnetic resonance-guided focused ultrasound (MRgFUS) has emerged as an established treatment for Essential Tremor (ET) and Tremor-Dominant Parkinson’s disease (TDPD). As a thermoablative procedure, MRgFUS creates permanent intracranial lesions to modulate pathological activity within tremor-related neural circuits. In both disorders, the most common target is the cerebello-thalamo-cortical tract (CTCT) at the level of the ventral intermediate nucleus (VIM), although TDPD additionally involves basal ganglia loop dysfunction^1^.

While outcomes in ET are generally favorable, tremor relapse after MRgFUS in TDPD remains substantial, affecting approximately 30-50% of patients^2^. The irreversible nature of lesioning introduces inherent uncertainty regarding optimal target locations. Prior studies linked relapse to smaller lesion volumes and younger age^2,3^, yet these descriptive factors do not explain why anatomically similar lesions yield divergent long-term outcomes.

The long-term benefits of MRgFUS thalamotomy for TDPD are variable, highlighting the need for a clearer definition of the connectivity profiles associated with a sustained clinical response. Although advantageous lesion sites were proposed^4,5^, increasing evidence suggests that durable tremor control depends less on a single anatomical coordinate than on effective modulation of disease-relevant networks^5^. Yet, the network-level determinants of tremor outcome following MRgFUS in TDPD remain poorly defined. In particular, it is unclear how the intrinsic connectivity of lesion locations relates to the degree and durability of tremor suppression across patients.

Here, we characterize the structural and functional connectivity profiles of MRgFUS lesion sites in relation to tremor improvement as a continuous measure, while additionally comparing patients with sustained benefit and those with relapse. Using normative connectomic mapping^6^, we assess how variation in the network embedding of thalamic lesions relates to clinical response. This network-based approach moves beyond single-target anatomy to examine how the connectivity architecture of lesion locations may shape therapeutic efficacy in Parkinsonian tremor.

## Materials and methods

### Patients and clinical testing

Twenty patients with TDPD, diagnosed according to International Parkinson and Movement Disorder Society criteria, underwent unilateral MRgFUS (ExAblate Neuro 4000, Insightec) targeting the VIM between December 2019 and April 2024. Targeting followed a standardized tractography-guided indirect approach^7^.

Clinical assessments were conducted at baseline and 6-month follow-up using the Unified Parkinson’s Disease Rating Scale^8^ (UPDRS)-III and Fahn-Tolosa-Marín Tremor scale^9^ (FTM). Tremor severity was defined by UPDRS-III items 3.15–3.18 and FTM parts A and B, with percentage tremor improvement at 6 months as the primary outcome. All evaluations were performed in a defined off-medication state (≥4h withdrawal). Levodopa Equivalent Daily Dosage (LEDD) was calculated using established conversion factors^10^.

Exclusion criteria were described previously^7^. All participants provided written informed consent (local ethic votum No. 314/18).

### Neuroimaging

MRI was performed on a 3T scanner (Achieva TX, Philips Healthcare, Best, The Netherlands) 2 days before, 3 days and 6 months after treatment. Imaging included high-resolution T1-weighted anatomical and diffusion-weighted sequences using standard clinical acquisition protocols described previously^7,11^ and in the Supplementary material.

Lesions were manually segmented on 6-month T1-weighted images in native space to capture the permanent coagulation necrosis. Right-sided lesions were flipped to the left hemisphere for anatomical comparability. Lesion masks were normalized to MNI152 space (2mm isotropic resolution.

### Functional and Structural connectivity mapping

Details of preprocessing and analytical procedures are reported in the Supplement material.

#### Functional Connectivity Analysis

Patients were classified as responder (>30% tremor improvement, n = 13) or relapser (<30%, n = 7), consistent with prior publications^2,12^. Lesion overlap maps were generated for each subgroup and thresholded (≥25% voxel overlap within each group)^4^ to define seed regions.

Seed-based functional connectivity was computed using normative resting-state functional MRI datasets from healthy controls (OASIS-3^13^, N = 301) and Parkinson patients (PPMI^14^, N = 256) respectively. Whole-brain voxel-wise connectivity maps were generated and Fisher Z-transformed. Differences between responder- and relapser-derived connectivity maps were assessed using general linear models (GLM).

#### Structural Connectivity Analysis

Structural connectivity was evaluated using probabilistic tractography in both, patient-specific diffusion MRI and a normative structural connectome sample (PPMI, N = 84). White matter streamlines which intersected with individual lesion masks were identified. To minimize spurious connections, only tracts with voxel overlap of >25% within the sample were retained.

Outcome-relevant pathways were identified by weighting tract involvement by individual tremor improvement and aggregating across patients. Statistical inference was performed using Wilcoxon-signed-rank test with permutation-based correction (5000 iterations).

#### Connectivity Fingerprinting and Predictive Modeling

Normative structural and functional connectome data (PPMI) were used within the Lead-Connectome framework^15^ (LeadDBSv2.5) to derive connectivity fingerprints. Individual lesion masks served as seeds to estimate whole-brain connectivity profiles.

Following the approach described by Horn et al^16^, connectivity maps from each lesion to every brain voxel within the functional and structural connectomes were correlated with contralateral tremor improvement across patients using Spearman’s rank correlation. This procedure yielded voxel-wise R-maps for both connectivity modalities, representing connectivity patterns associated with favorable clinical outcome.

We evaluated predictive validity using a leave-one-patient-out cross-validation (LOOCV) approach, whereby the optimal connectivity map was iteratively recomputed by excluding one patient at a time. The similarity between the held-out R-map and the optimal connectivity model was then used to estimate clinical improvement. Finally, positively associated cortical regions were structurally back-projected to the thalamus to identify connectivity-informed target locations^17^.

## Statistical Analysis

Demographic and clinical variables were analyzed using SPSS (Version 27, IBM Corp., 2020.). Longitudinal evaluation of outcome variables was conducted using mixed ANOVA with time and group factors.

Connectivity analyses and tractography were conducted with *FSL* and MATLAB-based toolboxes and are described in more detail in the Supplementary material.

## Results

### Patient Demographics

In the overall cohort, tremor severity contralateral to the treated VIM decreased significantly 6 months after MRgFUS. UPDRS-III tremor scores significantly improved by −41%, and FTM-A/B scores by −51% (Table 1). Tremor improvement was unrelated to lesion volume or age but correlated positively with disease duration (r = 0.71, *p* = 0.007), baseline tremor severity (UPDRS-III: r = 0.64, *p* = 0.02; FTM-A/B: r = 0.46, *p* = 0.040), and baseline LEDD (r = 0.72, *p* = 0.006).

**Table 1.**
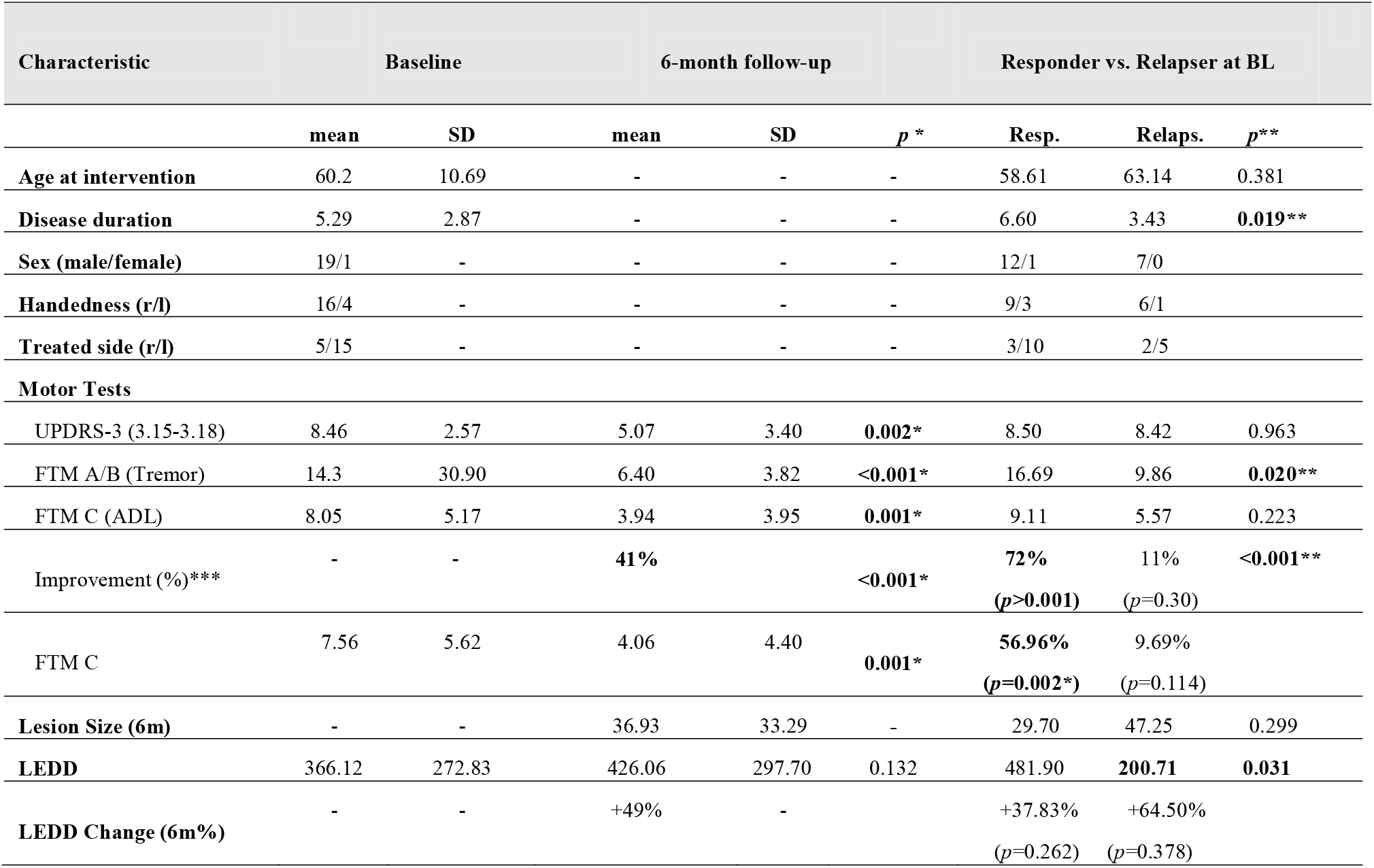
Patient Demographics and Clinical Characteristics. Patient demographics and clinical characteristics at baseline (BL) and 6-month follow-up, including comparison between responder and relapser subgroups at baseline. Mixed ANOVAs were used to assess changes from baseline to 6 months within the overall cohort, with significant differences indicated by *p*-values (<0.001) marked with * or ** when comparing responder and relapser. Improvement scores *** represent percentage change in tremor severity, quantified using the UPDRS-III tremor subscore (or FTM A/B when UPDRS-III was unavailable) with their significance testing BL vs. Follow-up.

| Characteristic | Baseline |  | 6-month follow-up |  | *p* * | Responder vs. Relapser at BL |  |  |
| --- | --- | --- | --- | --- | --- | --- | --- | --- |
|  | mean | SD | mean | SD |  | Resp. | Relaps. | *p* ** |
| Age at intervention | 60.2 | 10.69 | - | - | - | 58.61 | 63.14 | 0.381 |
| Disease duration | 5.29 | 2.87 | - | - | - | 6.60 | 3.43 | **0.019\*\*** |
| Sex (male/female) | 19/1 | - | - | - | - | 12/1 | 7/0 |  |
| Handedness (r/l) | 16/4 | - | - | - | - | 9/3 | 6/1 |  |
| Treated side (r/l) | 5/15 | - | - | - | - | 3/10 | 2/5 |  |
| **Motor Tests** |  |  |  |  |  |  |  |  |
| UPDRS-3 (3.15-3.18) | 8.46 | 2.57 | 5.07 | 3.40 | **0.002\*** | 8.50 | 8.42 | 0.963 |
| FTM A/B (Tremor) | 14.3 | 30.90 | 6.40 | 3.82 | **<0.001\*** | 16.69 | 9.86 | **0.020\*\*** |
| FTM C (ADL) | 8.05 | 5.17 | 3.94 | 3.95 | **0.001\*** | 9.11 | 5.57 | 0.223 |
| Improvement (%)*** | - | - | **41%** |  | **<0.001\*** | **72%**<br>(*p*>**0.001**) | 11%<br>(*p*=0.30) | **<0.001\*\*** |
| FTM C | 7.56 | 5.62 | 4.06 | 4.40 | **0.001\*** | **56.96%**<br>(*p*=**0.002\***) | 9.69%<br>(*p*=0.114) |  |
| Lesion Size (6m) | - | - | 36.93 | 33.29 | - | 29.70 | 47.25 | 0.299 |
| LEDD | 366.12 | 272.83 | 426.06 | 297.70 | 0.132 | 481.90 | **200.71** | **0.031** |
| LEDD Change (6m%) | - | - | +49% | - |  | +37.83%<br>(*p*=0.262) | +64.50%<br>(*p*=0.378) |  |

Responder (>30% improvement) exhibited M = −72% tremor reduction at 6 months, whereas relapser (<30%) showed non-significant M = −11% reduction (*p* = 0.30). Noteworthy, both groups did not differ significantly (*p* = 0.221) regarding their tremor improvement at 3 days (−95% vs. −79%; Supplementary material). Groups did not differ in age or lesion volume, but responder had longer disease duration and higher baseline FTM-A/B scores. Baseline LEDD differed between groups (*p* = 0.031) but not at follow-up, and medication remained unchanged.

### Functional Connectivity Profiles of Responder vs. Relapser

Thresholded lesion overlap maps of responder and relapser are shown in Figure 1a and served as seeds for subsequent connectivity analyses (Figure 1b-c). Although partially overlapping with the relapser map, responder lesions were located more medially at the VIM–ventral caudal nucleus (VC) border, aligning with our previously defined sweetspot^4^. In contrast, relapser lesions were positioned more laterally, extending toward the zona incerta and inferior ventro-oralis-posterior (VOp).

**Figure 1.**
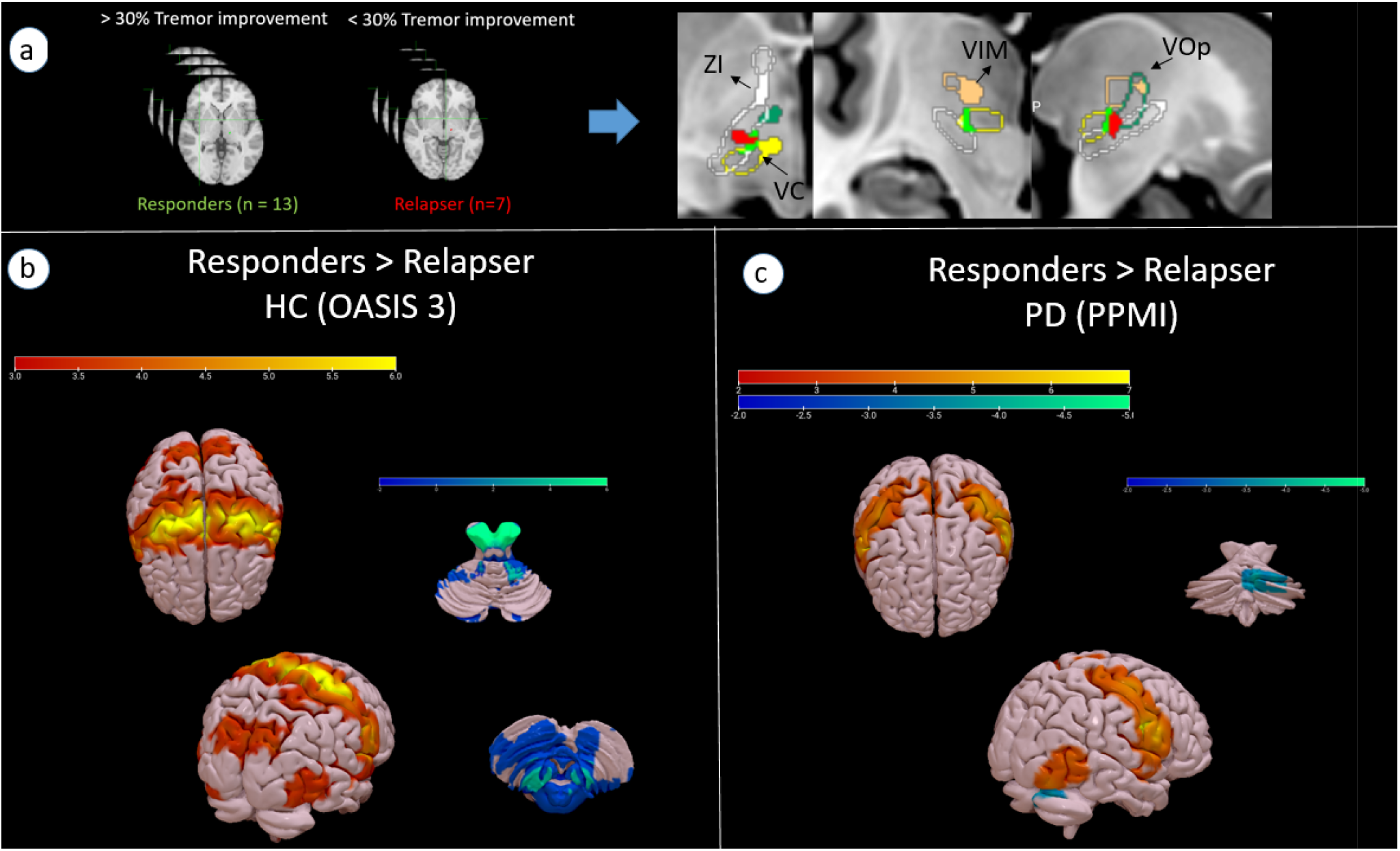
Functional Connectivity Profiles of Responder and Relapser. (**A**) Seed-based connectivity (SBC) analysis comparing responder and relapser lesion sites. Patients were classified as responder (>30% tremor improvement) or relapser (<30% tremor improvement). Lesion masks from each group were merged separately and thresholded at 25% voxel-wise agreement to define group-level seeds. SBC analyses were performed using two independent functional connectomes (**B**) the OASIS-3 database (n = 301, healthy controls) and (**C**) the PPMI database (n = 256, Parkinson’s disease patients). Group differences in connectivity were assessed using a paired *t*-test within a GLM. Regions showing stronger connectivity from responder compared with relapser lesion sites are displayed in warm colors, whereas regions with stronger connectivity from relapser compared with responder lesion sites are shown in cool colors.

GLM analysis of seed-based functional connectivity for OASIS-3 revealed distinct network embeddings. The responder seed showed stronger normative connectivity to bilateral motor-related cortical regions, including primary motor cortex (M1), postcentral gyrus, and supplementary motor area. Additional differences were observed in inferior frontal and occipito-visual regions, including V2 and the lateral occipital complex (Figure 1b). These findings were replicated using a PPMI normative connectome (Figure 1c), confirming stronger connectivity of responder lesions to Parkinson’s-specific motor territories.

Conversely, the relapser seed exhibited stronger connectivity to cerebellar regions. In the healthy connectome, this included bilateral Crus I and lobules IV-VI, and VIII, while the PPMI connectome showed increased connectivity to right Crus I and lobule VI.

### Structural connectivity of effective tremor control

Figure 2 depicts the tractography-based structural connectivity profile of lesion sites associated with the highest tremor improvement. At the subcortical level, significant streamlines consistently traversed the previously published sweetspot for TDPD tremor control^4^, located at the posteroinferior VIM along the VIM-VC boundary. From this subcortical convergence zone, streamlines extended superiorly through the posterior VOp to cortical motor regions, predominantly terminating in the M1 and S1.

**Figure 2.**
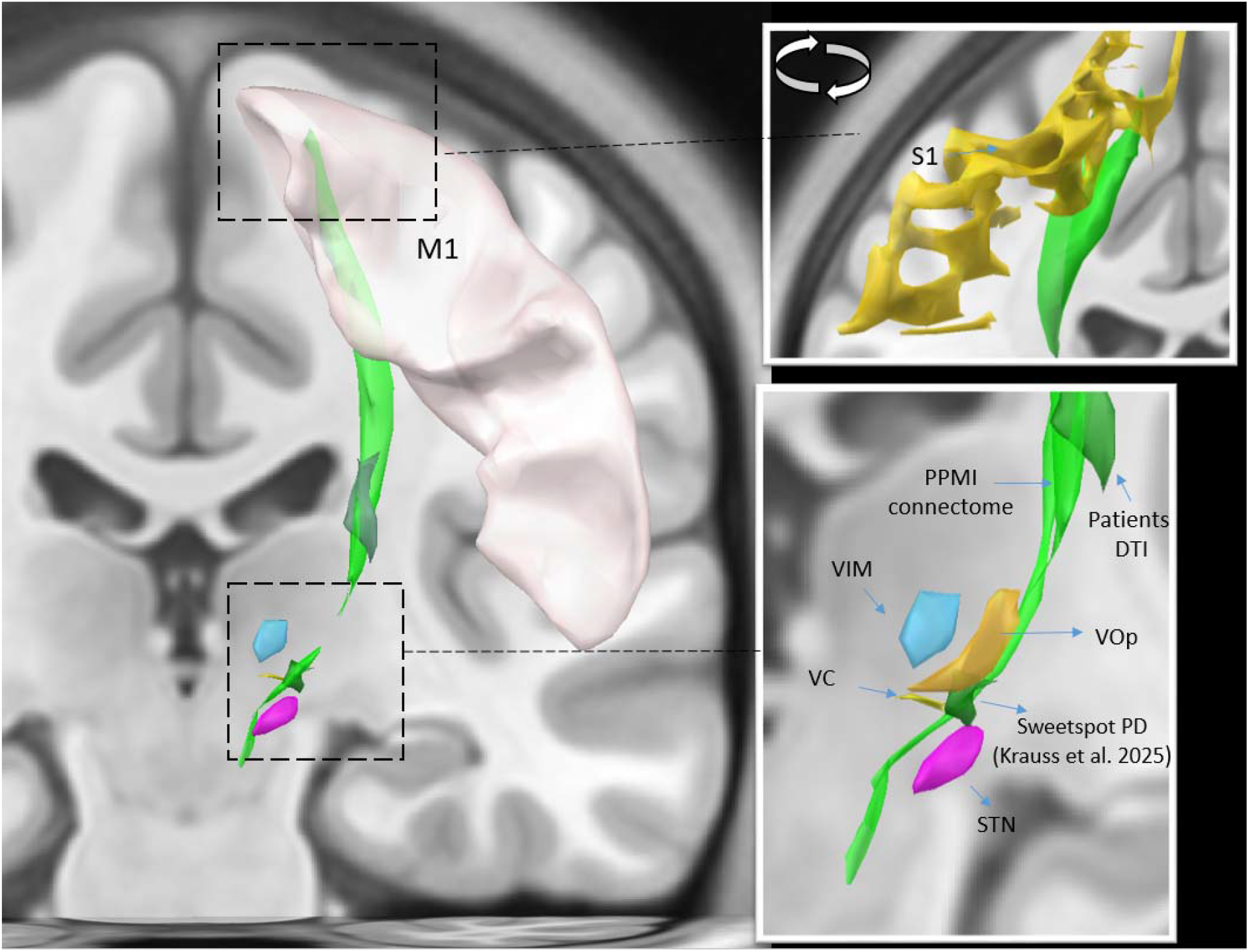
Optimal Structural Connectivity of Effective Tremor Control. Figure 2 depicts the structural connectivity profile of lesion sites associated with the highest tremor improvement, as derived from tractography analyses. At the subcortical level, significant streamlines consistently traversed the previously published sweet spot for PD tremor control^4^, located at the border between the VIM and the VC. From this subcortical convergence zone, streamlines extended superiorly along the VOp to cortical motor regions, predominantly terminating in the sensorimotor cortex. All anatomical regions are incorporated in the DISTAL atlas.

### Structural and Functional Connectivity Fingerprints for Optimal MRgFUS treatment

Structural connectivity analysis revealed that lesion sites whose streamlines preferentially connected to the S1 were associated with greater tremor improvement (Figure 3b). Among sensorimotor regions, S1 connectivity showed strongest positive association with clinical outcome, with streamlines further extending toward posterior cortical regions, including occipital areas. In contrast, lesion sites exhibiting stronger structural connectivity to regions anterior to M1 demonstrated a negative association with clinical improvement. Using LOOCV, the structural connectivity model showed significant internal predictive validity (r = 0.46, *p* = 0.04), indicating that the identified structural connectivity fingerprint reliably captured inter-individual differences in clinical response.

**Figure 3.**
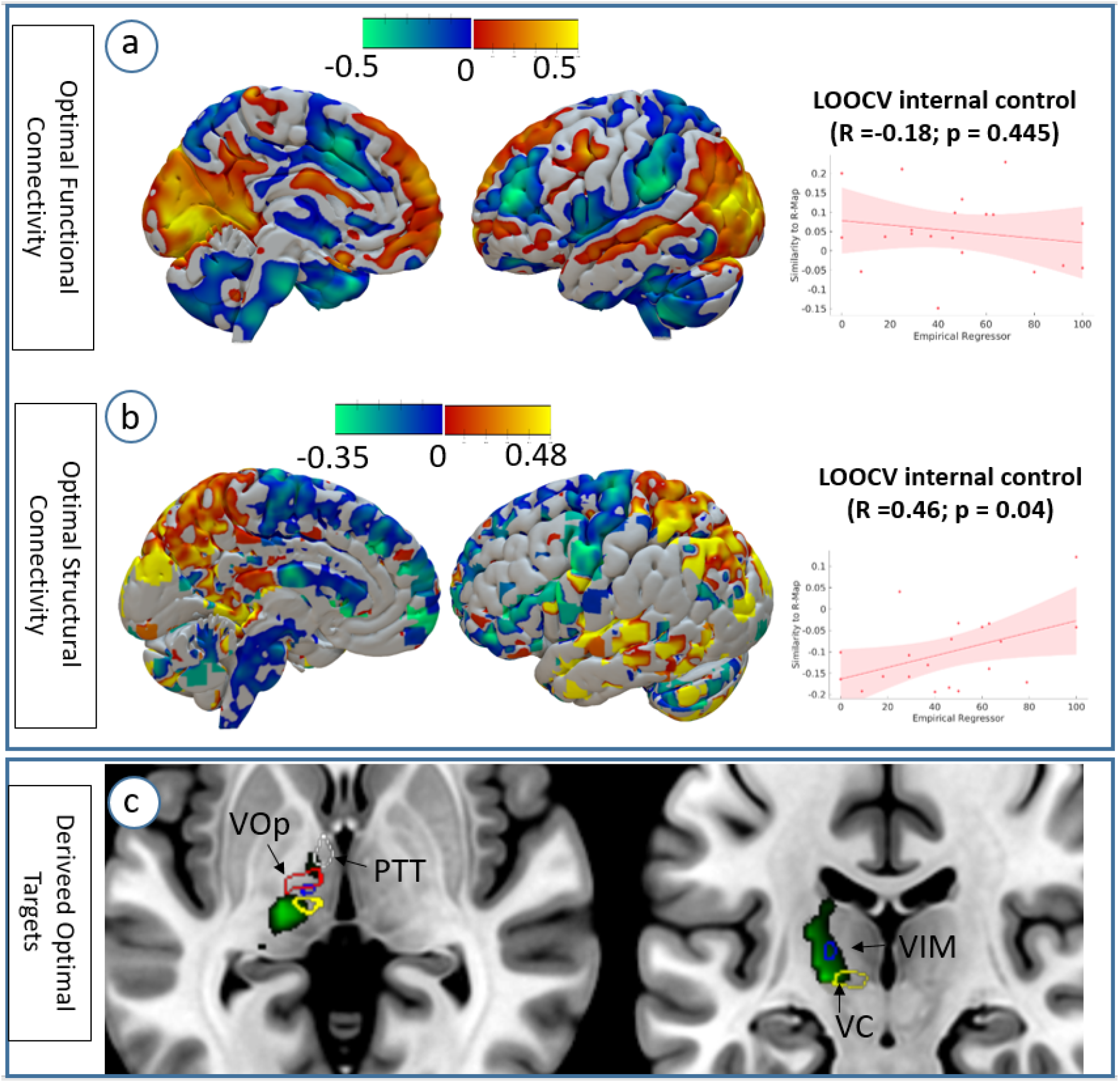
Optimal Functional and Structural Connectivity Fingerprints. Standardized z-maps and tractographies derived from normative PPMI functional (A) and structural (B) connectomes were correlated with percentage tremor improvement to identify connectivity patterns associated with clinical outcome (R-Maps). Regions whose connectivity to the lesion site correlated positively with tremor reduction are shown in warm colors, whereas regions associated with limited or absent clinical benefit are shown in cool colors. Predictive performance was internally validated using LOOCV, in which each subject was iteratively excluded as a test case while the model was trained on the remaining cohort. (C) Cortical regions demonstrating the strongest positive associations in the structural connectivity fingerprint were used as seeds for structural connectivity analysis using the thalamus as an output mask. This analysis identified connectivity-informed target regions at the VIM/VC border and within Forel’s field H1 as candidate loci associated with favorable clinical response.

Functional connectivity fingerprint analysis demonstrated a partially overlapping but less robust pattern. Lesion sites with stronger functional connectivity to M1 and occipital cortical regions were associated with greater tremor improvement (Figure 3a). However, in contrast to structural connectivity, the functional connectivity model failed internal validation through LOOCV (r = −0.18, *p* = 0.445).

Back-projection of positively associated cortical voxels onto the thalamus localized connectome-guided hotspots located primarily at the VIM–VC border, but also more anteriorly at the thalamic fasciculus (Forel’s field H1), where the PTT intersects the thalamus (Figure 3c).

## Discussion

We examined functional and structural connectivity hallmarks across the clinical improvement spectrum following MRgFUS thalamotomy for TDPD to define network correlates of durable tremor control. In contrast to prior reports^2^, lesion size and age were not predictive in our cohort. Instead, greater baseline tremor severity and longer disease duration were associated with larger tremor reduction, suggesting that patients with more advanced tremor phenotypes may derive greater benefit from MRgFUS.

At the network level, sustained improvement was associated with lesion sites showing stronger functional connectivity to M1, S1, and SMA, as well as to inferior frontal and occipital cortices. In contrast, lesion sites with poorer outcomes showed greater connectivity with cerebellar motor and associative regions, particularly contralesional Crus I and lobules IV-VI and VIII.

Streamlines associated with better outcomes projected posteriorly toward S1, with M1 delineating an anterior functional boundary beyond which outcomes worsened. Whole-brain structural fingerprints consistently emphasized S1 and M1, underscoring sensorimotor thalamo-cortical projections as a key therapeutic axis.

### Functional Connectivity Profiles of Responder vs Relapser

Our findings support the triangular interface of VIM-VC-VOp as an effective target, while suggesting that long-term efficacy may be better explained by connectivity profiles than by anatomical location alone.

Lesion sites associated with sustained tremor improvement demonstrated stronger functional connectivity to motor regions, including M1, SMA, and S1, as well as to occipital cortices. A comparable motor connectivity pattern has been described for VIM-DBS in ET^17^, and the cortical distribution observed here overlaps partially with the recently proposed and validated somato-cognitive action network (SCAN)^18,19^. Given that SCAN-related hyperconnectivity is characteristic of PD and not observed in ET, differential network engagement may explain why anatomically similar lesion sites yield distinct outcomes in ET and TDPD^18,19^. Notably, the close spatial proximity of responder and relapser sites underscores that even minor deviations of targets and corresponding connectivity profiles may translate into clinically meaningful differences.

In contrast, responder lesions exhibited relatively lower functional connectivity to motor and associative regions within the cerebellum. Although the VIM receives cerebellar input via the dentate nucleus^20^, the lower cerebellar connectivity of the responder relatively to the relapse lesion site, functional cerebellar modulation may be less important for durable tremor control. These findings remain observational but contribute to the ongoing discussion regarding the extent to which tremor generation can be attributed to cerebellar mechanisms alone^21^.

### Structural target network for effective TDPD tremor treatment

Lesions associated with superior outcomes intersected our previously published sweetspot^4^ and engaged streamlines projecting posteriorly along VOp towards M1 and S1. This pattern is broadly consistent with the tractographic observations of Cheyou et al.^5^, although projections in our cohort did not prominently involve premotor territories.

Although aforementioned group localized the optimal target more anterior at the VIM/VOp border, the streamlines identified here likely converge on overlapping pathways, including components of the CTCT and possibly the pallido-thalamic tract (PTT) - both implicated in tremor suppression in TDPD. At the same time, evidence that DBS may be less effective at the VIM/VOp border than near the VC, where tremor cells are more frequently encountered^22^, highlights that the precise structural substrate of therapeutic benefit remains unresolved.

Within current pathophysiological frameworks, including the “dimmer-switch” model^1^, basal ganglia circuits are thought to initiate pathological oscillations, whereas tremor expression is mediated through CTCT projections with M1 as a central cortical node. Our findings align with this model by emphasizing thalamo-cortical output towards M1 and S1 as a structural correlate of effective tremor control, consistent with prior VIM-DBS studies^23^, while underscoring the need to further delineate the relative contributions of individual fiber systems.

### Connectomic signatures of tremor improvement in TDPD

Across modalities, favorable outcomes were associated with lesion sites preferentially connected to S1, whereas connectivity extending anterior than M1/SMA border predicted poorer results. Although functional cross-validation did not reach statistical significance and should be interpreted cautiously, the convergence of structural and functional findings underscores the relevance of the sensorimotor cortex within the tremor network.

This pattern aligns with connectivity-based DBS findings^17^ and electrophysiological evidence implicating postcentral regions in tremor-related oscillatory dynamics^24–26^, and is anatomically supported by established thalamocortical projections from VIM and VC to S1^1^.

A less intuitive component of the improvement-related network involves visual areas. Structural and functional alterations of occipital cortex have been reported in tremor disorders^27; 7^ and may carry predictive relevance^28^. Their precise mechanistic role remains uncertain but likely reflects broader network-level modulation rather than direct motor effects.

## Clinical Relevance

The present findings delineate connectivity features associated with durable MRgFUS outcomes in TDPD, highlighting network characteristics that may inform patient-specific targeting in a population where relapse remains common. We identified connectivity patterns linked to sustained benefit, confirmed our previously reported sweetspot^4^, and - through inversion of the optimal structural fingerprint - localized an overlapping region with a slight inferoposterior extension that reproduces the desired connectivity profile aligning with the VIM-DBS target proposed by Milosevic et al (2018)^29^. Notably, the inverted map extends toward regions typically lesioned in PTT ablations also targeted in the stepwise dual-lesion approach described by Chen et al.^30^, suggesting that VIM- and PTT-directed interventions may represent complementary strategies for modulating a shared tremor network.

Beyond anatomical targeting, these findings support a shift toward connectivity-guided, individualized treatment planning. Connectomic signatures may serve as imaging biomarkers to predict treatment response and refine lesion placement on a patient-specific basis. Integrating demographic and clinical variables that may influence cortical connectivity (e.g. disease duration, tremor severity, LEDD) may further improve patient selection and targeting. Continued validation in independent cohorts will be essential to translate these insights into personalized MRgFUS strategies.

## Limitations

This study is limited by its retrospective design, precluding causal inference and limiting control over unmeasured confounders. The relatively small sample size further constrains generalizability, despite the usage of large normative datasets and the generally standardized nature of MRgFUS procedures with careful patient selection. Replication in larger, prospective cohorts with external validation is therefore warranted.

Connectivity analyses relied in part on normative rather than disease-specific connectomes. Although normative datasets may not fully capture individual or disorder-specific network alterations, prior studies^6^ suggest broadly comparable results between normative and disease-specific approaches. To reduce false-positive findings, we applied stringent statistical thresholds and, where available, integrated normative connectivity with patient-specific lesion and connectivity data to enhance internal validity.

## Data availability

The data that support the findings of this study are available on reasonable request from the corresponding author

## Supplementary material

Supplementary material is available at *Brain* online.

